# Incidence of occult cancer in patients diagnosed with adenocarcinoma in situ by cervical punch biopsy or endocervical curettage

**DOI:** 10.1101/2024.01.11.24301143

**Authors:** Byung-Su Kwon, Dong Hoon Suh, Myong Cheol Lim, Kidong Kim, Chae Hyeong Lee, Hyun Hoon Chung, Yong-Beom Kim

## Abstract

**Objectives:** To investigate the incidence of occult cancer in patients diagnosed with adenocarcinoma in situ through punch biopsy or endocervical curettage.

**Methods:** We examined the clinicopathological characteristics of patients diagnosed with adenocarcinoma in situ using punch biopsy or endocervical curettage at four institutes in the Republic of Korea from 2003 to 2011. We investigated the incidence of occult cancer in conization or hysterectomy specimens and whether the occult cancer was detected through conization.

**Results:** Twenty patients diagnosed with adenocarcinoma in situ through punch biopsy or endocervical curettage were enrolled. Six out of the 20 patients had occult cervical cancer, and four out of the six patients had cervical cancer at or greater than stage 1A1 with lymphovascular space invasion. Two out of four patients were diagnosed with invasive cervical cancer through conization and underwent radical hysterectomy without adjuvant radiotherapy. The other two patients underwent extrafascial hysterectomy without preoperative conization and received adjuvant radiotherapy.

**Conclusion:** A significant number of patients diagnosed with adenocarcinoma in situ through punch biopsy or endocervical curettage have occult cancer. Conization should be performed to identify occult cancer and optimize the extent of surgery.

## Introduction

Adenocarcinoma in situ (AIS) is a precursor of cervical adenocarcinoma and its incidence is increasing. This is supported by an analysis of 9 population-based cancer registries in the United States, which revealed an increasing prevalence of AIS in young women [1]. The standard treatment for AIS is not conization but extrafascial hysterectomy because AIS can exist as skipped lesions and be located in the deep endocervix [2]. However, for women who want to preserve their fertility, conization alone can be considered as an alternative treatment [3].

The management of AIS diagnosed using punch biopsy or endocervical curettage (ECC) is unclear because previous studies have only addressed the management of AIS diagnosed using conization. To our knowledge, no study has addressed the optimal management of AIS diagnosed using punch biopsy or ECC.

For example, unless a patient does not need to preserve fertility, she has two options: conization followed by hysterectomy or extrafascial hysterectomy without preoperative conization. The first option may allow for an accurate diagnosis before hysterectomy but requires two separate operations, whereas the second option carries the risk of suboptimal surgery for occult cancer. If the incidence of occult cancer is significant, performing conization before hysterectomy may be a more reasonable approach than performing extrafascial hysterectomy without preoperative conization.

The objective was to investigate the incidence of occult cancer in patients diagnosed with AIS through punch biopsy or ECC.

## Methods

After obtaining approval from the Institutional Review Board (IRB) at each institution (B-1203-148-106), clinicopathological variables of patients diagnosed with AIS by punch biopsy or ECC at four hospitals in the Republic of Korea between 2003 and 2011 were collected through a review of medical records. Acquisition of informed consent was waived by the IRB. The collected variables included age, results of Papanicolaou (Pap) smear and human papillomavirus (HPV) test, pathological results of conization and hysterectomy, type of hysterectomy, and adjuvant therapy.

We examined the incidence of occult cancer, the incidence of cervical cancer at or greater than stage 1A1 with lymphovascular space invasion (LVSI), and whether these occult cancers were detected through conization.

## Results

Twenty patients were included in this study, and their characteristics are summarized in Table 1. The median age of the patients was 42 years, and the most frequent result of the Pap smear test was atypical glandular cells. High-risk HPV was detected in the majority of patients. Only four patients underwent conization followed by a hysterectomy.

**Table 1.** Patient characteristics (n = 20)

|  |  |
| --- | --- |
| Age, median (range), years | 42 (25–78) |
| Pap smear results, n |  |
| Unknown | 2 |
| Normal | 2 |
| Abnormal, no specific result | 1 |
| ASC-US | 2 |
| LSIL | 2 |
| AGC | 6 |
| ASC-H | 2 |
| HSIL | 1 |
| AIS | 1 |
| Adenocarcinoma | 1 |
| HPV test, n |  |
| Not done | 5 |
| Done | 15 |
| High-risk HPV positive | 12 |
| High-risk HPV negative | 3 |
| Treatment, n |  |
| Conization alone | 7 |
| Conization followed by hysterectomy | 4 |
| Hysterectomy without preoperative conization | 9 |

Pap, Papanicolaou; ASC-US, atypical squamous cells of undetermined significance; LSIL, low-grade squamous intraepithelial lesion; AGC, atypical glandular cells; ASC-H, atypical squamous cells cannot exclude high-grade squamous intraepithelial lesion; HSIL, high-grade squamous intraepithelial lesion; AIS, adenocarcinoma in situ; HPV, human papillomavirus

Finally, six out of 20 patients were diagnosed with invasive cervical cancer. Of these, two patients were diagnosed with cervical cancer stage 1A1 without LVSI and four patients had cervical cancer at or greater than stage 1A1 with LVSI. Two out of the four patients with cervical cancer at or greater than stage 1A1 with LVSI underwent conization and received radical hysterectomy without adjuvant radiotherapy. However, the other two patients underwent extrafascial hysterectomy without preoperative conization and received postoperative radiotherapy (Table 2).

**Table 2.** Summary of cases with cervical cancer at or greater than stage 1A1 with LVSI.

| Age<br>(years) | Pap test finding | HPV | Treatment | Final diagnosis |
| --- | --- | --- | --- | --- |
| 30s | AGC | Negative | Conization followed<br>by hysterectomy | Cervical cancer 1A2 |
| 40s | AGC | 18 | Conization followed<br>by hysterectomy | Cervical cancer 1B1 |
| 40s | ASC-H | 16 | Hysterectomy<br>without preoperative<br>conization | Cervical cancer 1B1 |
| 50s | Adenocarcinoma | Negative | Hysterectomy<br>without preoperative<br>conization | Cervical cancer 1B1 |

LVSI, lymphovascular space invasion; Pap, Papanicolaou; AGC, atypical glandular cells; ASC-H, atypical squamous cells cannot exclude high-grade squamous intraepithelial lesion; HPV, human papillomavirus

## Discussion

We found that 30% of patients diagnosed with AIS using punch biopsy or ECC had occult cancer, and two-thirds of them had disease equal to or more advanced than stage 1A1 with LVSI. Therefore, performing an extrafascial hysterectomy without preoperative conization is considered undertreatment, which can result in adjuvant radiotherapy in a significant number of patients.

In addition, our study suggested that conization is sensitive in detecting occult cancer in patients diagnosed with AIS using punch biopsy or ECC. Specifically, occult cancer was identified in all patients with cervical cancer at or more advanced than stage 1A1 with LVSI undergoing conization.

Previous studies have suggested that conization is necessary for diagnosing AIS. For example, a study including 24 cases of AIS reported that the diagnosis of AIS using colposcopy and cervical biopsy was difficult [4]. Another study on AIS reported that conization was necessary in most cases to detect invasive cancer [5]. Similarly, a study including 21 cases with AIS reported that invasive cancer cannot be excluded by punch biopsy [6]. However, previous studies did not clearly describe the number of occult cancers detected by conization. They also have a limitation in that most patients were diagnosed with AIS by conization rather than punch biopsy or ECC.

The strengths of our study are the clear descriptions of the number of patients who underwent punch biopsy or ECC, conization, and hysterectomy. The weaknesses of our study are the small number of enrolled patients and the retrospective design.

In conclusion, a substantial number of patients diagnosed with AIS through punch biopsy or ECC have occult cancer. Conization should be performed to identify occult cancer and optimize the extent of surgery.

## Data Availability

All data produced in the present study are available upon reasonable request to the authors

## Ethical Approval

All procedures performed in studies involving human participants were conducted in accordance with the ethical standards of the institutional and/or national research committee, as well as the 1964 Helsinki Declaration and its subsequent amendments or equivalent ethical standards.

